# BODY MASS INDEX TRAJECTORY BEFORE DEATH IN COMMUNITY-DWELLING OLDER ADULTS. POPULATION-BASED COHORT STUDY

**DOI:** 10.1101/2021.05.21.21257579

**Authors:** Hari V Pai, Martin C Gulliford

## Abstract

**Background and objective:** Both low and high body mass index (BMI) have been associated with greater mortality in older adults. This study evaluated the trajectory of BMI in the final years of life.

**Design:** Population-based cohort study.

**Setting:** Community-dwelling adults in the English Longitudinal Study of Ageing between 1998 and 2012.

**Measurements:** Body mass index, years before death and all-cause mortality. Analyses were adjusted for age, gender, educational level, housing tenure and social class.

**Results:** Data were analysed for 16,924 participants with 31,857 BMI records; mean age at study start, 61.6 (SD 10.9) years; mean BMI, 27.5 (4.7) Kg/m^2^. There were 3,686 participants (4,794 BMI records) who died and 13,238 participants (27,063 BMI records) who were alive at last follow-up. Mean BMI increased with age to 60-69 years but then declined, but the age-related decline was more rapid in decedents. At ages 80-89 years, mean BMI in decedents was 26.1 (4.7) compared with 27.1 (4.4) Kg/m^2^ in survivors. After adjusting for age and covariates, mean BMI declined in the five years before death. From 9 to 5 years before death or end of study, adjusted mean BMI was 0.51 (95% confidence interval 0.24 to 0.78) Kg/m^2^ lower for decedents than survivors; and from four to zero years before death, 1.55 (1.26 to 1.84) Kg/m^2^ lower in decedents.

**Conclusions:** In community-dwelling older adults, mean body mass index enters an accelerating decline during five years before death. Reverse causation may account for the association of lower BMI with mortality.

## INTRODUCTION

The recent increase in obesity has accentuated the development of multiple morbidity at middle- and older-ages,(1) focusing attention on the association of body mass index with mortality. (2) Meta-analyses of cohort studies suggest that there is strong association between BMI and all-cause mortality (3-5), but there is evidence of ‘U’ or ‘J’-shaped relationship with either low or high body mass index being associated with greater mortality than normal body weight or overweight. (6-9) Similar patterns of association have been reported for systolic blood pressure (10) and total cholesterol values,(11) with either low or high blood pressure or total cholesterol being associated with higher mortality. Longitudinal analysis of blood pressure and cholesterol values recorded into electronic health records found evidence that both blood pressure and total cholesterol values decline towards the end of life. (12, 13) This suggested a possible role of reverse causation in explaining the associations of low blood pressure or low total cholesterol with mortality. We hypothesised that body mass index may show a declining trajectory towards the end of life. We analysed data from a cohort of older adults, the English Longitudinal Study of Ageing (ELSA),(14) in order to evaluate changes in BMI values in the period before death.

## METHODS

### Study design and participants

A retrospective cohort study was conducted using the English Longitudinal Study of Ageing (ELSA). The initial ELSA cohort included community-dwelling adults aged 50 and over, and their younger partners, drawn from households that responded to the Health Survey for England (HSE) in 1998, 1999 and 2001 (wave 0). Participants were 16,924 participants aged 40-90 years with data for BMI at wave 0. Patients aged less than 40 years were excluded, as were participants aged more than 90 years because their exact ages were not available for analysis. A flow chart for participant and data selection is shown in Supplementary Figure 1.

Participants gave their consent to participate in the study and ethical approval was granted from the London Multicentre Research Ethics Committee.

### Measures

Measurements of height and weight were recorded by nurse fieldworkers at waves 0, 2, 4 and 6. Standing height was measured using a portable stadiometer without shoes. One measurement was taken with participants stretching with the head in the Frankfort plane. Weight was measured in light clothing using portable electronic scales.(15) One BMI value of <10 Kg/m^2^ was omitted. Mortality was all-cause mortality up to February 2012, the latest date when the ELSA study mortality data were updated from the National Health Service Central Data Registry (NHSCR) records. (16) Covariates included age at measurement and the number of years before death. For decedents, years till death was calculated as the difference between the year of death recorded in the ELSA dataset and the year of measurement. For survivors, years till end of study was calculated as 2012 (the last year of mortality follow-up) minus the year of measurement. Because follow-up was conducted in waves, only selected values of years till end of study were possible for survivors. Covariate data from wave 0 included gender, highest educational qualifications (categories: not applicable; Degree or equivalent; Higher education below degree; A Level or equivalent; GCE O Level or equivalent; Other; Foreign qualification; No qualification), housing tenure (categories: Owner; Buying with mortgage; part rent and part mortgage; Rented; live rent free; other and not applicable) and social class (categories: Professional; Managerial technical; Skilled non-manual; Skilled manual, Semi-skilled manual; Unskilled manual; Armed forces; other and not applicable).

### Statistical analysis

Mean BMI values were evaluated by age and year till death. Multiple linear regression was used to model repeated BMI values over time, including a random effect at the participant-level using the ‘xtreg’ command in Stata version 14.(17) There was strong evidence that the association of BMI with age and years till death differed by mortality status. Models were fitted separately for men and women who died (decedents) and those who were alive at the end of follow-up (survivors). Age and years till death were fitted as continuous variables, together with quadratic terms to allow for non-linearity, adjusting for education, housing tenure and social class. Predicted BMI from adjusted models were compared with mean BMI values and plotted using the ‘ggplot2’ package(18) in the R program.(19)

## RESULTS

The cohort included 31,857 BMI records for 16,924 participants. The mean age at wave 0 was 61.6 (SD 10.9) years with, mean BMI, 27.5 (4.7) Kg/m^2^. There were 3,686 participants, with 4,794 BMI records, who died; and 13,238 participants, with 27,063 BMI records who did not die at end of follow-up. Table 1 shows the distribution of mean BMI by gender, age-group and years till death or end of study. Mean BMI was generally similar for men and women, but the standard deviation for BMI was greater in women consistent with the greater prevalence of severe obesity in women. In survivors, mean BMI increased from 27.1 Kg/m^2^ at age 40-44 years, to 28.1 Kg/m^2^ between 55 to 74 years, before declining to 26.6 Kg/m^2^ at age 85 to 90 years. A similar pattern was observed in decedents, but the highest BMI was observed between 45 and 64 years and the mean BMI in the oldest age-group was lower at 25.6 Kg/m^2^. In survivors, mean BMI tended to increase over time from 27.4 Kg/m^2^ at 14 years before the end-of-study to 28.2 Kg/m^2^ at the end-of-study. In decedents, mean values were less stable because of the smaller number of observations. However, mean BMI declined in the last five years of life from 27.5 Kg/m^2^ five years before death to 26.0 Kg/m^2^ in the last year of life.

**Table 1:** Body mass index by gender, age-group and years till death or end of study. Figures are mean (SD) body mass index.

|  |  | Survivors |  | Decedents |  |
| --- | --- | --- | --- | --- | --- |
|  |  | Freq. | Mean BMI (SD) | Freq. | Mean BMI (SD) |
| **Gender** | Men | 11,657 | 27.8 (4.1) | 2,496 | 27.2 (4.5) |
|  | Women | 15,406 | 27.9 (5.3) | 2,298 | 27.0 (5.3) |
| **Age at measurement (years)** | 40-44 | 411 | 27.1 (5.8) | 7 | 27.1 (4.5) |
|  | 45-54 | 5,294 | 27.5 (4.8) | 283 | 28.1 (6.3) |
|  | 55-64 | 9,436 | 28.1 (5.0) | 732 | 28.1 (5.4) |
|  | 65-74 | 7,724 | 28.1 (4.7) | 1,534 | 27.5 (5.0) |
|  | 75-84 | 3,677 | 27.7 (4.5) | 1,801 | 26.6 (4.5) |
|  | 85-90 | 521 | 26.6 (4.4) | 437 | 25.6 (3.9) |
| **Years till death or end of study** | 0 | 3,228 | 28.2 (5.1) | 150 | 26.0 (4.9) |
|  | -1 |  | - | 385 | 26.4 (5.0) |
|  | -2 |  | - | 443 | 26.7 (4.6) |
|  | -3 | 2,483 | 28.3 (5.3) | 448 | 27.0 (5.2) |
|  | -4 | 2,314 | 28.3 (5.0) | 416 | 26.8 (4.9) |
|  | -5 |  | - | 425 | 27.5 (4.9) |
|  | -6 |  | - | 471 | 27.4 (5.2) |
|  | -7 | 3,212 | 28.0 (4.8) | 421 | 26.9 (5.0) |
|  | -8 | 2,588 | 27.9 (4.8) | 350 | 27.5 (4.9) |
|  | -9 |  | - | 318 | 27.2 (4.7) |
|  | -10 |  | - | 333 | 27.2 (4.6) |
|  | -11 | 5,261 | 27.8 (4.7) | 242 | 27.7 (4.7) |
|  | -12 |  |  | 191 | 28.1 (5.5) |
|  | -13 | 2,621 | 27.4 (4.6) | 165 | 27.5 (4.7) |
|  | -14 | 5,356 | 27.4 (4.5) | 36 | 28.0 (4.4) |

Figure 1 (left panel) shows mean BMI values (points) and predicted BMI values adjusted for covariates (lines) by single year of age for survivors (blue) and decedents (red). BMI increased up to approximately age 65 years. Over the age of 70 years, mean BMI declined with each increasing year of age, with the decline being more rapid in decedents than survivors. In survivors, points were generally close to predicted values; in decedents, while points were close to predicted values at older ages, at young ages where there were fewer deaths, there were outlying values of either high BMI values (more evident in men) or low BMI values (more evident in women).

**Figure 1:**
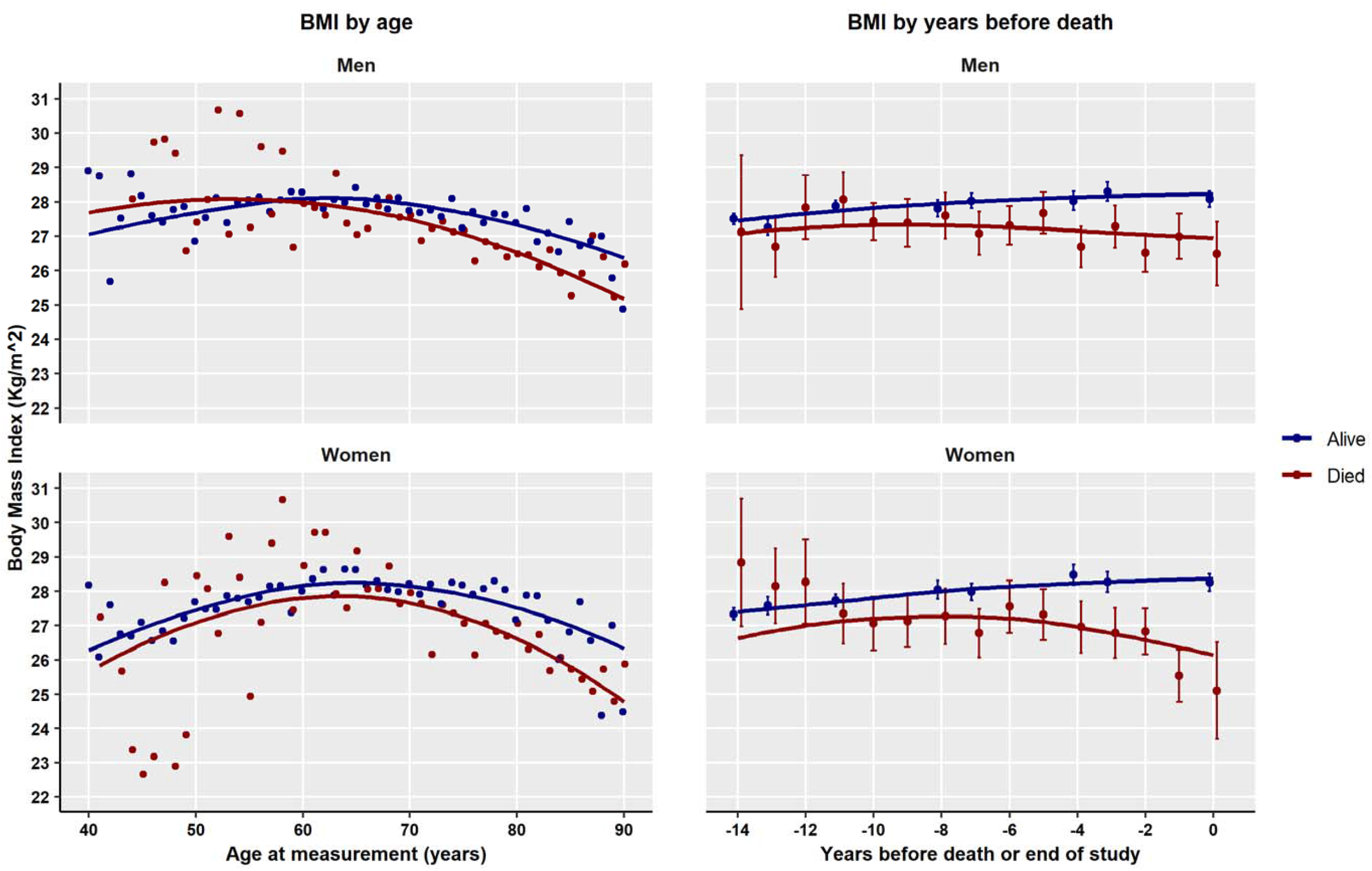
Mean BMI by age (left panel) and mean BMI by years before death or end of study (right panel) for decedents (red) and survivors (blue). Points are mean BMI values, lines are predicted values from model adjusted for age, years till death, education, housing tenure and social class. Error bars are 95% confidence intervals.

Figure 1 (right panel) shows mean BMI values (95% confidence intervals) by year before death for decedents or end-of-study for survivors, together with predicted values from the adjusted model. Differences between data points and fitted lines may in part be accounted for by covariate adjustment. While BMI remained stable or increased in survivors, there was evidence of declining BMI towards the end of life in decedents. Between zero and four years before death, after adjusting for covariates, BMI was 1.55 (95% confidence interval 1.26 to 1.84) Kg/m^2^ lower in decedents than survivors; between five and nine years before death, BMI was 0.51 (0.24 to 0.78) Kg/m^2^ lower in decedents than survivors.

## DISCUSSION

### Main findings

BMI enters a decline over the age of 65 to 70 years and this decline is earlier and more rapid in decedents than survivors. After adjusting for age and covariates, BMI was found to show an accelerated decline in the last five years of life, and this was not evident at the end of study in survivors. Diehr et al.(20) observed that the processes of ageing and dying may be difficult to distinguish. While body mass index decline may a feature of ageing, accelerated loss of body mass may result from pathological processes leading to mortality.

Consequently, associations of lower body mass index with mortality (9) are likely to be explained by ‘reverse’ causation, where mortality risk is the predictor of BMI.

### Strengths and limitations

The present analyses drew on data for a well-established, population-based cohort study with well-described measurement techniques.(14) The main limitations of the study are the limited response rate of 67% at wave 0 and attrition at subsequent waves, (14) leading to a sample that might not be representative over time. Mortality follow-up was restricted to the period up to 2012. Survivors might be misclassified if they died soon after the end of follow-up. Body mass index records were limited to those participants who participated in measurements during years in which follow-up waves of data collection were conducted.

### Conclusions and Implications

In the WHO classification of obesity, the mean BMI value for the cohort was consistent with ‘overweight’ rather than ‘healthy’ body weight. For older adults, BMI occupies an ambiguous position with respect to healthy ageing: higher BMI is a risk factor for long-term conditions, including type II diabetes and hypertension; but BMI loss may also be associated with elevated mortality probability. In order to evaluate this, research and clinical assessment might not only consider body weight but also give attention to understanding the prognostic significance of additional indicators including body fat distribution and body composition.

## Data Availability

Data for the English Longitudinal Study of Ageing may be accessed through the UK Data Archive.

## Acknowledgement

This work was conducted as part of a medical student elective at King’s College London, GKT School of Medicine.

## REFERENCES

1. Booth HP, Prevost AT, Gulliford MC. Impact of body mass index on prevalence of multimorbidity in primary care: cohort study. Family Practice. 2014;31(1):38–43.

2. Di Angelantonio E, Bhupathiraju Sh N, Wormser D, Gao P, Kaptoge S, et al. Body-mass index and all-cause mortality: individual-participant-data meta-analysis of 239 prospective studies in four continents. Lancet. 2016;388(10046):776–86.

3. Aune D, Sen A, Prasad M, Norat T, Janszky I, Tonstad S, et al. BMI and all cause mortality: systematic review and non-linear dose-response meta-analysis of 230 cohort studies with 3.74 million deaths among 30.3 million participants. BMJ. 2016;353:i2156.

4. Berrington de Gonzalez A, Hartge P, Cerhan JR, Flint AJ, Hannan L, MacInnis RJ, et al. Body-mass index and mortality among 1.46 million white adults. N Engl J Med. 2010;363(23):2211–9.

5. Whitlock G, Lewington S, Sherliker P, Clarke R, Emberson J, et al. Body-mass index and cause-specific mortality in 900 000 adults: collaborative analyses of 57 prospective studies. Lancet. 2009;373(9669):1083–96.

6. Ng TP, Jin A, Chow KY, Feng L, Nyunt MSZ, Yap KB. Age-dependent relationships between body mass index and mortality: Singapore longitudinal ageing study. PLoS One. 2017;12(7):e0180818.

7. Yuan JM, Ross RK, Gao YT, Yu MC. Body weight and mortality: a prospective evaluation in a cohort of middle-aged men in Shanghai, China. Int J Epidemiol. 1998;27(5):824–32.

8. Jee SH, Sull JW, Park J, Lee SY, Ohrr H, Guallar E, et al. Body-mass index and mortality in Korean men and women. N Engl J Med. 2006;355(8):779–87.

9. Bhaskaran K, dos-Santos-Silva I, Leon DA, Douglas IJ, Smeeth L. Association of BMI with overall and cause-specific mortality: a population-based cohort study of 3.6 million adults in the UK. The Lancet Diabetes & Endocrinology. 2018;6(12):944–953. doi: 10.1016/S2213-8587(18)30288-2.

10. Guo Z, Viitanen M, Winblad B. Low blood pressure and five-year mortality in a Stockholm cohort of the very old: possible confounding by cognitive impairment and other factors. Am J Public Health. 1997;87(4):623–8.

11. Petersen LK, Christensen K, Kragstrup J. Lipid-lowering treatment to the end? A review of observational studies and RCTs on cholesterol and mortality in 80+-year olds. Age and Ageing. 2010;39(6):674–80.

12. Ravindrarajah R, Hazra NC, Hamada S, Charlton J, Jackson SHD, Dregan A, et al. Systolic Blood Pressure Trajectory, Frailty, and All-Cause Mortality >80 Years of Age: Cohort Study Using Electronic Health Records. Circulation. 2017;135(24):2357–68.

13. Charlton J, Ravindrarajah R, Hamada S, Jackson SH, Gulliford MC. Trajectory of Total Cholesterol in the Last Years of Life Over Age 80 Years: Cohort Study of 99,758 Participants. J Gerontol A Biol Sci Med Sci. 2018;73(8):1083–9.

14. Steptoe A, Breeze E, Banks J, Nazroo J. Cohort Profile: The English Longitudinal Study of Ageing. Int J Epidemiol. 2013;42(6):1640–8.

15. Zaninotto P, Lassale C. Socioeconomic trajectories of body mass index and waist circumference: results from the English Longitudinal Study of Ageing. BMJ Open. 2019;9(4):e025309.

16. Dregan A, Ravindrarajah R, Hazra N, Hamada S, Jackson SHD, Gulliford MC. Longitudinal Trends in Hypertension Management and Mortality Among Octogenarians: Prospective Cohort Study. Hypertension. 2016;68:97–105

17. Stata Corp. Stata Statistical Software: Release 14. College Station, TX: StataCorp LP; 2015.

18. Wickham H. ggplot2: Elegant graphics for data analysis. Heidelberg: Springer; 2009.

19. R Core Team. R: A language and environment for statistical computing. Vienna, Austria: R Foundation for Statistical Computing; 2016.

20. Diehr P, Williamson J, Burke GL, Psaty BM. The aging and dying processes and the health of older adults. J Clin Epidemiol. 2002;55(3):269–78.

